# Diagnosis value of SARS-CoV-2 antigen/antibody combined testing using rapid diagnostic tests at hospital admission

**DOI:** 10.1101/2020.09.19.20197855

**Authors:** Nicolas Veyrenche, Karine Bolloré, Amandine Pisoni, Anne-Sophie Bedin, Anne-Marie Mondain, Jacques Ducos, Michel Segondy, Brigitte Montes, Patrick Pastor, David Morquin, Alain Makinson, Vincent Le Moing, Philippe Van de Perre, Vincent Foulongne, Edouard Tuaillon

## Abstract

**Objectives:** The implementation of rapid diagnostic tests (RDTs) may enhance the efficiency of SARS-CoV-2 testing, as RDTs are widely accessible and easy to use. The aim of this study was to evaluate the performance of a diagnosis strategy based on a combination of antigen and IgM/IgG serological RDTs.

**Methods:** Plasma and nasopharyngeal samples were collected between 14 March and 11 April 2020 at hospital admission from 45 patients with RT-PCR confirmed COVID-19 and 20 negative controls. SARS-CoV-2 antigen (Ag) was assessed in nasopharyngeal swabs using the Coris Respi-Strip. For IgM/IgG detection, SureScreen Diagnostics and Szybio Biotech RDTs were used in addition to laboratory assays (Abbott Alinity i SARS-CoV-2 IgG and Theradiag COVID-19 IgM ELISA).

**Results:** Using the Ag RDT, 13 out of 45 (29.0%) specimens tested positive, the sensitivity was 87.0% for Cycle Threshold (CT) values ≤ 25 and 0% for CT values > 25. IgG detection was associated with high CT values and the amount of time after the onset of symptoms. The profile of isolated IgM on RDTs was more frequently observed during the first and second week after the onset of symptoms. The combination of Ag and IgM/IgG RDTs enabled the detection of up to 84.0% of COVID-19 confirmed cases at hospital admission.

**Conclusion:** Antigen and antibody-based RDTs showed suboptimal performances when used alone. However when used in combination, they are able to identify most COVID-19 patients admitted in an emergency department.

## I. Introduction

Reported for the first time in December 2019, COVID-19 has become a major public health concern worldwide. Currently, clinical management of COVID-19 is mainly based on the prevention of transmission, viral tests, and supportive care. Wide access to SARS-CoV-2 testing is one of the keys to protecting populations. To be efficient, diagnostic assays must be accessible in different settings, ranging from the hospital to the community level, and from low to high incomes countries ^1^. Reverse transcriptase polymerase chain reaction (RT-PCR) for the detection of SARS-CoV-2 RNA in upper and lower respiratory tract specimens (nasopharyngeal swab, throat swab, and sputum) is the gold standard to confirm COVID-19 ^2^. RT-PCR tests have an overall sensitivity estimated around 70.0% in nasopharyngeal sampling^3^ with a high specificity. RT-PCR is ideal for the diagnosis of COVID-19 during the first week after the onset of symptoms because the viral load is high during this period. Beyond day 14, when the viral load is low or undetectable, the performance of RT-PCR diminishes ^4^. In such situations, serological tests may help to confirm a COVID-19 diagnosis in individuals with a high clinical suspicion but who tested negative for SARS-CoV-2 RNA.

Both nucleic acid tests and automated serological tests require sample collection, transportation, and laboratory analysis, leading to a delayed response and limiting the efficiency of SARS-CoV-2 testing strategies. Furthermore, insufficient access to nearby laboratory facilities is a major concern in intermediate and low-income countries. A diagnostic test is characterized not only by its analytical performance, mainly estimated by sensitivity (Se) and specificity (Sp) but also by its overall accessibility ^5^. The implementation of rapid diagnostic tests (RDTs) in the diagnosis of COVID-19 could have significant benefits by enhancing the efficiency of large testing strategies ^6^. RTDs are useful devices that facilitate testing outside of laboratory settings, a capability needed for hard to reach populations ^7^. In addition, RTDs deliver results in a shorter amount of time than RT-PCR. This time saving is important for the identification, isolation and provision of appropriate clinical care to patients with COVID-19. RTDs also reduce overloads in emergency departments ^8^.

COVID-19 RDTs are based on the detection of either SARS-CoV-2 antigen in respiratory specimens or anti-SARS-CoV-2 antibodies in whole blood or plasma/serum. Experience with RDTs used to detect antigens from other respiratory viruses in respiratory samples suggests that the sensitivity of these tests is lower than that of nucleic acid tests, ranging from 34.0% to 80.0% ^9^. Recent publications have confirmed that COVID-19 RDTs are considerably less sensitive than molecular tests, and may therefore generate false negative results ^10^. Antigen detection is mainly dependant on the viral concentration, hence most specimens with high viral concentrations test positive for antigen ^11–13^. In contrast to RDTs based on Ag detection, RDTs that detects anti-SARS-CoV-2 antibodies are widely available, and a very large number have been approved by the FDA and CE. Although weak or absent humoral responses have been reported, especially in mild and moderate forms of COVID-19, most patients develops an antibody response within the first two weeks of COVID-19 ^14–16^. Serologic assays also can be useful in conjunction with molecular assays for the clinical assessment of persons who present themselves for testing long after the onset of symptoms. While RDTs might constitute a simple screening method, they have shown limitations in the early phase of acute infections due to the time required for an antibody response. Consequently, COVID-19 diagnosis based on IgM and IgG detection is often delayed to the second phase of the disease, when some opportunities for therapy and prevention of SARS-CoV-2 transmission already have been lost. Due to the overall performance of the tests, WHO and FDA do not recommend the use of Ag or antibody-detecting RDTs as the sole basis for the diagnosis of infection, but are encouraging research studies to establish their usefulness. Diagnostic algorithms based on Ag plus antibodies detection using RDTs need to be compared to the molecular techniques which currently are the gold standard for COVID-19 diagnosis ^7^. The aim of this study was to evaluate the performance of a combination of antigen and serological RDTs to diagnose COVID-19 in hospitalized patients who tested positive for SARS-CoV-2 RNA using RT-PCR.

## II. Material and Methods

Plasma and nasopharyngeal samples were collected from patients admitted in Montpellier University hospitals between 14 March and 11 April 2020 who tested positive for SARS-CoV-2 RNA. Patient characteristics are detailed in Table 1. The estimated date of the onset of symptoms was recorded and ranged from 1 day to 20 days before hospital admission. The severity of COVID-19 was defined by WHO guidelines ^17^. Controls consisted of samples collected in the pre-COVID-19 period (2017-2018) in patients and stored at −80°C until used (DC-2015-2473). The cohort received an institutional ethics committee approval (CPP Ile de France III, n°2020-A00935−34; ClinicalTrials.gov Identifier: NCT04347850).

**Table 1:** Patient characteristics

|  | **Ct ≤ 25\*** | **25 < Ct < 35\*** | **Ct ≥ 35\*** | **Controls\*\*** |
| --- | --- | --- | --- | --- |
| Number of patients | 15 | 15 | 15 | 20 |
| Age (median, SD) | 66 (48-84) | 63 (50-76) | 58 (49-67) | 64 (35-93) |
| Sex ratio M/F | 9/6 | 13/2 | 10/5 | 10/10 |
| Days post-infection (median, SD) | 7 (4-10) | 8 (4-12) | 11 (7-15) | - |
| Severe COVID-19 | 9 | 8 | 9 | - |

\* Cycle Threshold (CT) values recorded by RT-PCR methods Allplex™ 2019-nCoV Assay Seegene.
\*\* Controls consisted of samples collected in the pre-COVID-19 period (2017-2018).
All serological and antigen assays were performed in strict accordance with the manufacturer's instructions.

### RT-PCR method

The Allplex™ 2019-nCoV Assay (Seegene, Seoul, South Korea) was used as the reference method to confirm SARS-CoV-2 infection. This RT-PCR simultaneously amplifies three different genes: the SARS-CoV-2 RNA-dependent RNA polymerase (RdRP) gene in the Cal Red 610 channel, the SARS-CoV-2 nucleocapsid (N) gene in the Quasar 670 channel, and the Sarbecovirus envelope (E) gene in the FAM channel. The result represents the positive result for the E gene of Sarbecovirus, the RdRP gene and the N gene of COVID-19, respectively. Nasopharyngeal samples were tested prospectively within a few hours after collection and without any cooling or freezing step. Swabs were collected in various transport media (eSwab™ COPAN Amies 1 ml, ∑-Transwab® liquid Amies, viral transport medium tube VTM-M 2.0ml). A 200µl nasopharyngeal sample was added in 200 µl Tissue Lysis Buffer (ATL) to inactivate the virus. These 400µl of sample were extracted and amplified by multiplex real time RT-PCR. COVID-19 confirmed-subjects were grouped according to the average value of the Cycle Threshold (CT), CT ≤ 25, 25 < CT < 35 and CT ≥ 35.

### Laboratory IgG and IgM immunoassays

Plasma samples were tested using the SARS-CoV-2 IgG immunoassay on the Alinity i system (Abbott, Illinois, USA). The SARS-CoV-2 assay is a chemiluminescent microparticule immunoassay (CMIA) intended for the qualitative detection of SARS-CoV-2 nucleoprotein IgG. The cut-off value for a positive result was defined by the manufacturer’s instructions: a ratio<1.4 calculated index (S/C) is considered negative and a ratio ≥ 1.4 is considered positive. Anti-SARS-CoV-2 IgM were not assessed by the Alinity i platform since the assay was not available at the time of the study.

IgM directed against SARS-CoV-2 protein S were detected using the ELISA COVID-19 THERA02 IgM assays (Theradiag, Marne la Vallée, France). The IgM positive cut-off is ratio≥1. All tests were performed according to the manufacturer’s instructions.

### Rapid diagnostic tests (RDTs)

The Coris COVID-19 Ag Respi-Strip (BioConcept®, Gembloux, Belgium), was used to test Ag in nasopharyngeal specimens. The assay is based on the detection of SARS-CoV-2 antigens in nasopharyngeal samples. This lateral flow assay uses colloidal gold nanoparticles sensitized with monoclonal antibodies directed against highly conserved SARS-CoV-2 nucleoprotein antigens.

Two SARS-CoV-2 lateral flow assays were evaluated. These RDTs use a chromatographic immunoassay format and are dedicated to the qualitative detection of IgG and IgM antibodies directed against SARS-CoV-2 in human whole blood, serum and plasma. The SureScreen Diagnostics Ltd (Derby, United Kingdom) COVID-19 IgG/IgM Rapid Test requires 10µl of sample collected and 80µl of buffer. The Szybio Biotech Joint Stock Co., Ltd. (Wuhan, China) SARS-CoV-2 IgM/IgG Antibody Assay Kit requires 10µl of serum, plasma or 20µl of whole blood sample and 60µl of buffer. During testing, the specimen reacts with SARS-CoV-2 antigen-coated particles in the test cassette. In the presence of a control signal, any signal visible in the IgM and/or IgG position at 15 minutes on the test line, even a weak one, must be interpreted as positive.

### Statistical analyses

The experimental data were summarized by number and percentage for categorical variable, *i.e*., positive and negative results. The analysis considers 45 patients; for each one we have a nasopharyngeal sample and a plasma sample collected on the same day. The 45 nasopharyngeal samples were positive for SARS-CoV-2 by RT-PCR. The controls consisted of 20 nasopharyngeal and plasma samples collected in the pre-COVID-19 period (2017-2018). COVID-19 confirmed patients were grouped in three categories according to RT-PCR values: CT ≤ 25, 25 < CT < 35 and CT ≥ 35. Analyses were performed using GraphPad Prism 8.0 (GraphPad Prism Software Inc., San Diego, California).

## III. Results

### RT-PCR CT results and SARS-CoV-2 serological results using laboratory assays

CT values were compared with the amount of time following the onset of symptoms. The two parameters were moderately correlated (R^2^= 0.3151, Fig.1).

**Figure 1.**
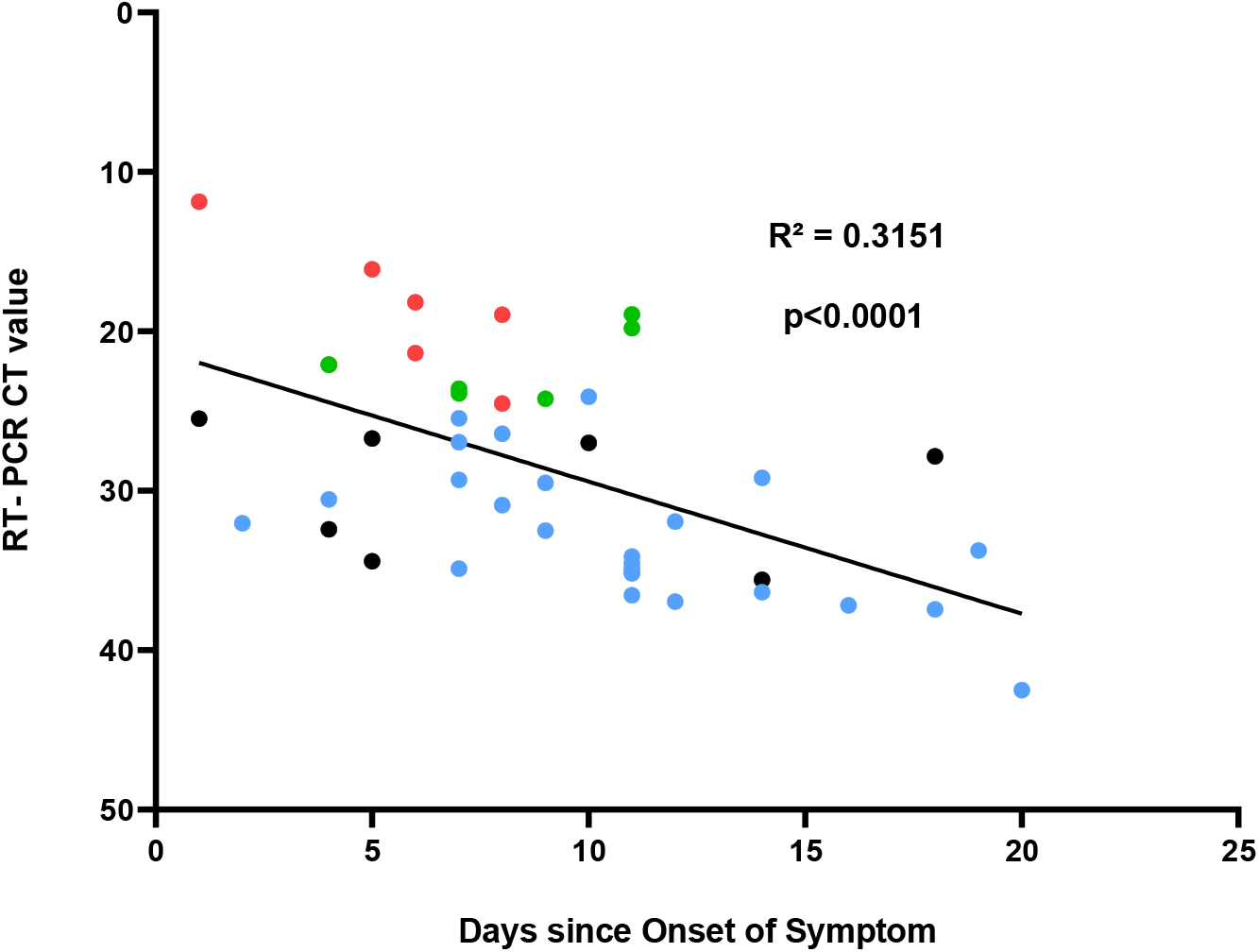
Correlation between RT-PCR CT values and the time delay from symptom onset. Cycle threshold (CT) values recorded by RT-PCR methods Allplex™ 2019-nCoV Assay Seegene. Each point corresponds to one patient and the color represents the RDT result. Red points correspond to patients who tested positive using the Coris RDT and negative with SureScreen RDT. Green points correspond to patients who tested positive with Coris and SureScreen RDTs. Blue points correspond to patients who tested positive using SureScreen RDT and negative with Coris RDT. Black points correspond to patients who tested negative with Coris and SureScreen RDTs.

The presence of anti-SARS-CoV-2 IgM and IgG was retrospectively assessed on plasma samples collected at admission in the emergency department using anti-SARS-CoV-2 IgG CMIA and IgM ELISA (Sup. Tables. S1. A and B).

A total of 24 out of 45 COVID-19 confirmed patients tested positive for IgG using the Abbott assay, resulting in a sensitivity (95% CI) of 53.3% (38.8 – 67.9) (Sup. Tables. S1. A and B). All but one patient tested two weeks after the onset of symptoms tested positive for IgG using the Abbott assay (Fig. 2). Signal to cut-off values were moderately correlated with the amount of time after the onset of symptoms (R^2^ = 0.3139, Fig. 2). CT values were higher in patients who tested positive for IgG than in IgG negative patients (median (IQR) = 33.1 (27.0 – 36.2); versus 25.5 (22.1 – 32.0), respectively (p=0.0073, Fig. 3). The IgG Abbott test had a sensitivity of 33.3% (9.2 – 56.8) for a CT value ≤ 25 at hospital admission, reaching 80.0% (59.8 – 100.0) for a CT value ≥ 35.

**Figure 2.**
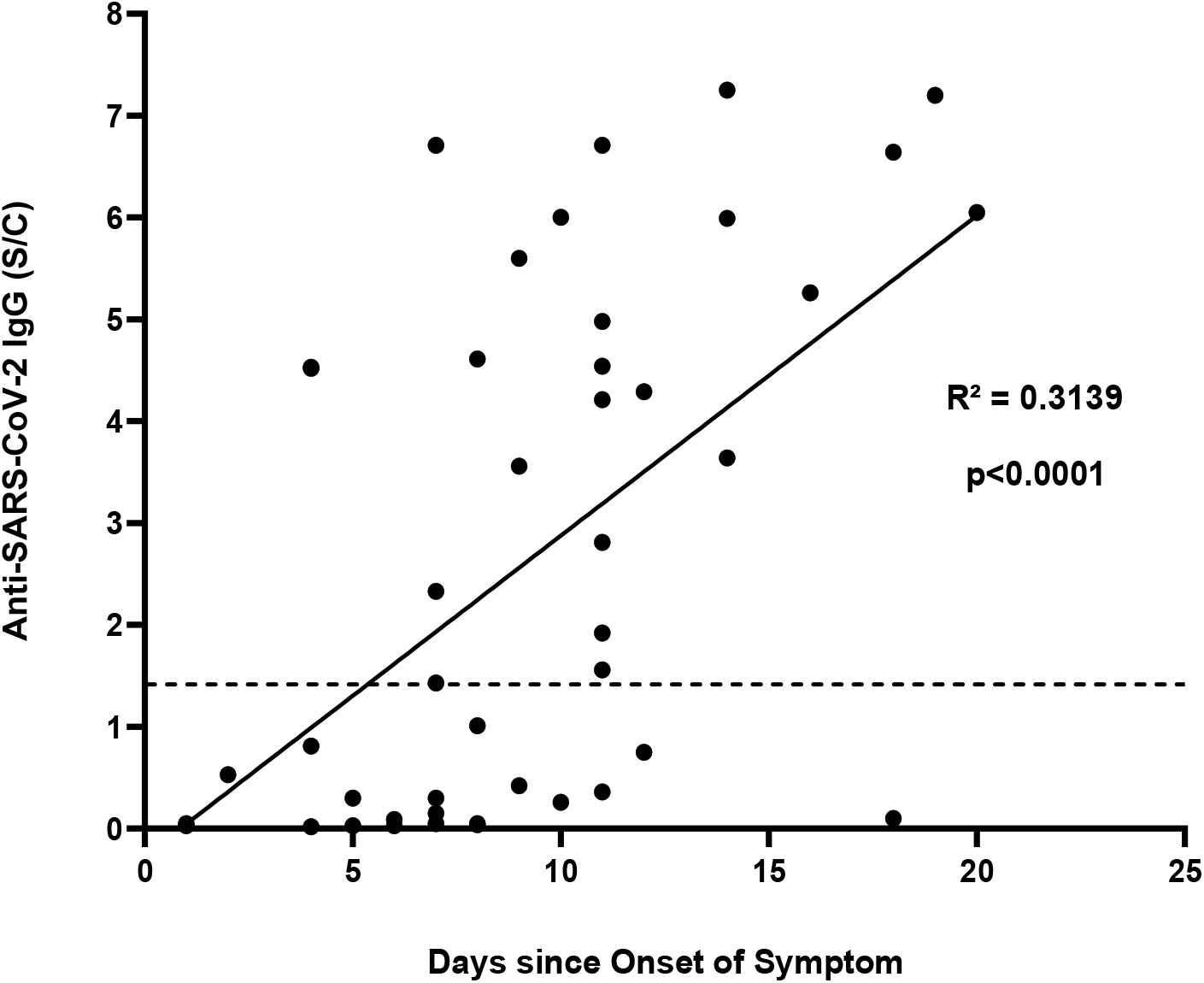
Correlation between Anti-SARS-CoV-2 IgG level and the time delay from symptom onset. Anti-SARS-CoV-2 IgG serology was performed on the Alinity i Abbott automated system and index of the signal to control values (S/C) were reported. The dotted line indicates the positive threshold (≥ 1.4).

**Figure 3.**
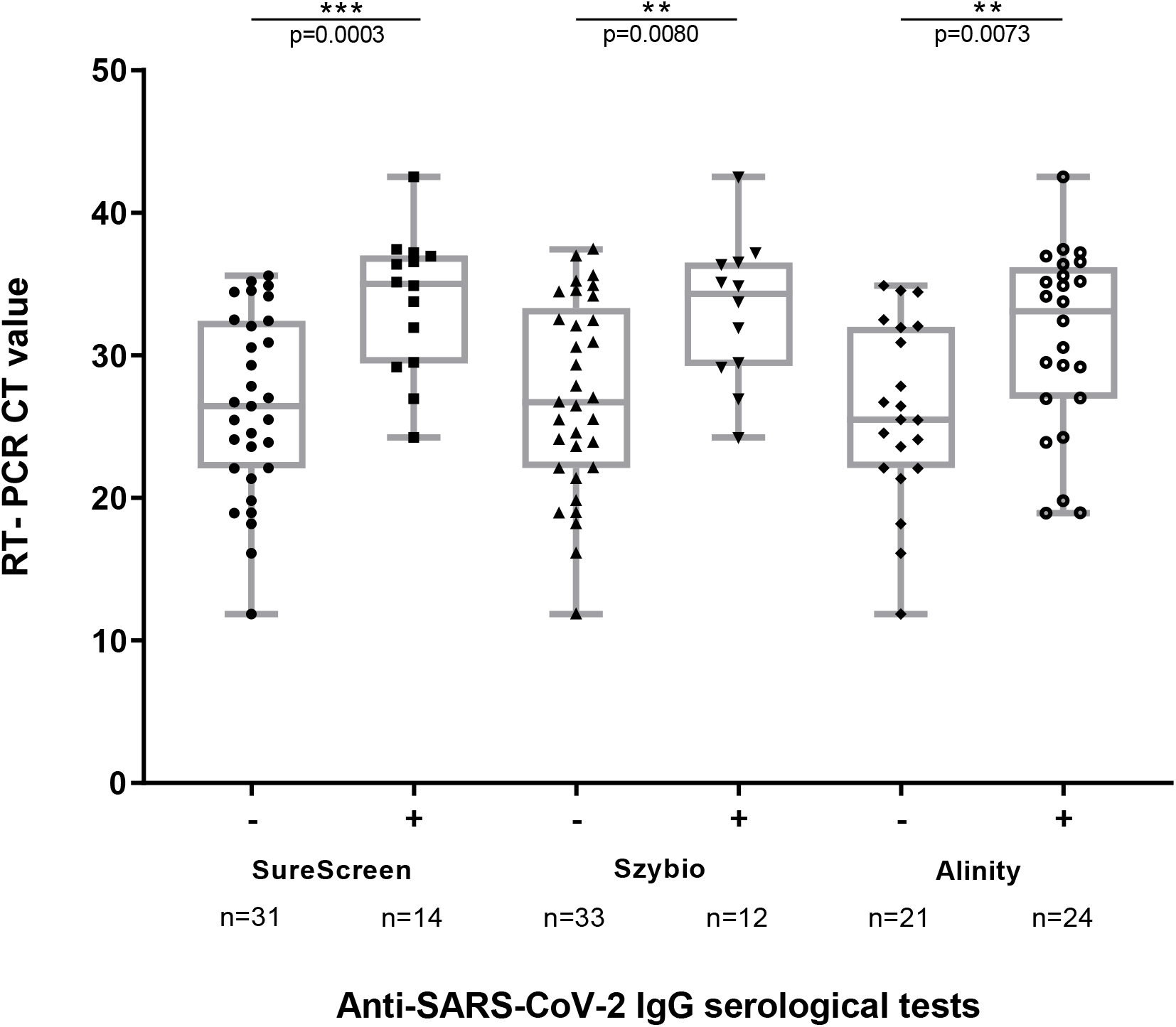
Anti-SARS-CoV-2 IgG detection using rapid serological diagnosis tests and chemoluminescence immunoassay according to RT-PCR CT values. Cycle threshold (CT) values recorded by RT-PCR methods Allplex™ 2019-nCoV Assay Seegene. The boxes represent interquartile ranges with the horizontal line indicating the median CT value and the whiskers showing minimal and maximal CT values. The *p* value was calculated using the Mann-Whitney U test, and compares the distribution of CT values in serological diagnosis tests with positive and negative results.

IgM directed against the S protein were detected in 24 specimens using the Theradiag ELISA, sensitivity (95% CI): 53.3% (38.8 – 67.9) (Sup. Tables. S1. A and B). Theradiag ELISA IgM ratios were not correlated with the amount of time after the onset of symptoms (R^2^= 0.0413) (data not shown). The CT values were higher in patients who tested positive for IgM, 32.8% (25.7 – 35.2) versus 26.7% (20.6 – 32.2) in IgM negative patients (p=0.0209, Fig. 4). The Theradiag IgM ELISA had a sensitivity (95% CI) of 33.3 % (9.2 – 56.8) for a CT value ≤ 25 at hospital admission, reaching 80.0% (59.8 – 100.0) for a CT value ≥ 35. Furthermore, sensitivity was 50.0% (29.1 – 70.9) when the estimated time since the onset of symptoms was ≤ 7 days, reaching 67.0% (36.3 – 97.7) ≥ 14 days (Sup. Tables S1. A and B).

**Figure 4.**
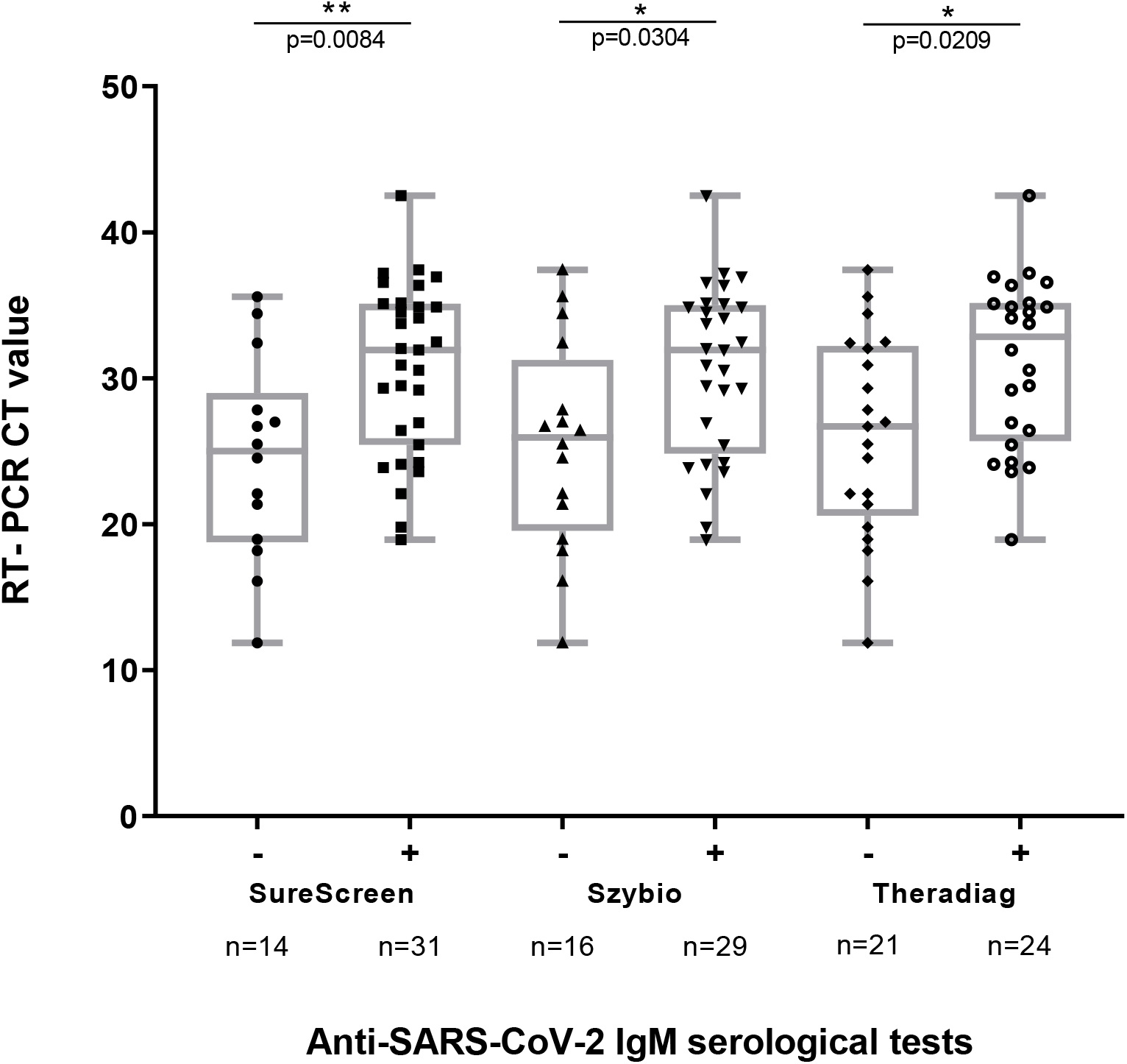
Anti-SARS-CoV-2 IgM detection using rapid serological diagnosis tests and ELISA according to RT-PCR CT values. Cycle threshold (CT) values recorded by RT-PCR methods Allplex™ 2019-nCoV Assay Seegene. The boxes represent interquartile ranges with the horizontal line indicating the median CT value and the whiskers showing minimal and maximal CT values. The *p* value was calculated using the Mann-Whitney U test, and compares the distribution of CT values in serological diagnosis tests with positive and negative results.

### SARS-CoV-2 antigen and IgM/IgG testing using RDTs

Nasopharyngeal and plasma samples then were tested for SARS-CoV-2 Ag and IgM/IgG using RDTs. All results of antigen and serologic RDTs were considered valid based on a visible control band, although the intensity of the band was weak for some specimens. Clinical samples collected from COVID-19 negative patients were used to assess the specificities of the RDTs. All controls samples tested negative for SARS-CoV-2 antigen and antibodies regardless of the assay (specificity=100.0%).

Among 45 nasopharyngeal samples collected in confirmed Covid-19 patients, 13 specimens tested positive using the Ag RDT, resulting in a sensitivity (95% CI) of 29.0% (15.7 – 42.3) (Fig.1 and Sup. Table S1. A). The median CT value was lower in Ag positive samples compared to negative ones (median (IQR) = 21.4 (18.6 – 23.8) versus 32.5 (28.2 – 35.2), p<0.0001, Fig. 5). The Ag RDT had a sensitivity (95% CI) of 87.0% (70.0 – 100.0) for CT values≤ 25 and 0% for CT values > 25 (Sup. Table 1.A). The sensitivity was 41.0% (20.4 – 61.6) in samples collected during the first week after the onset of symptoms, 29.0% (5.2 – 52.8) during the second week, and 0% after 14 days or more (Sup. Table S1. B).

**Figure 5.**
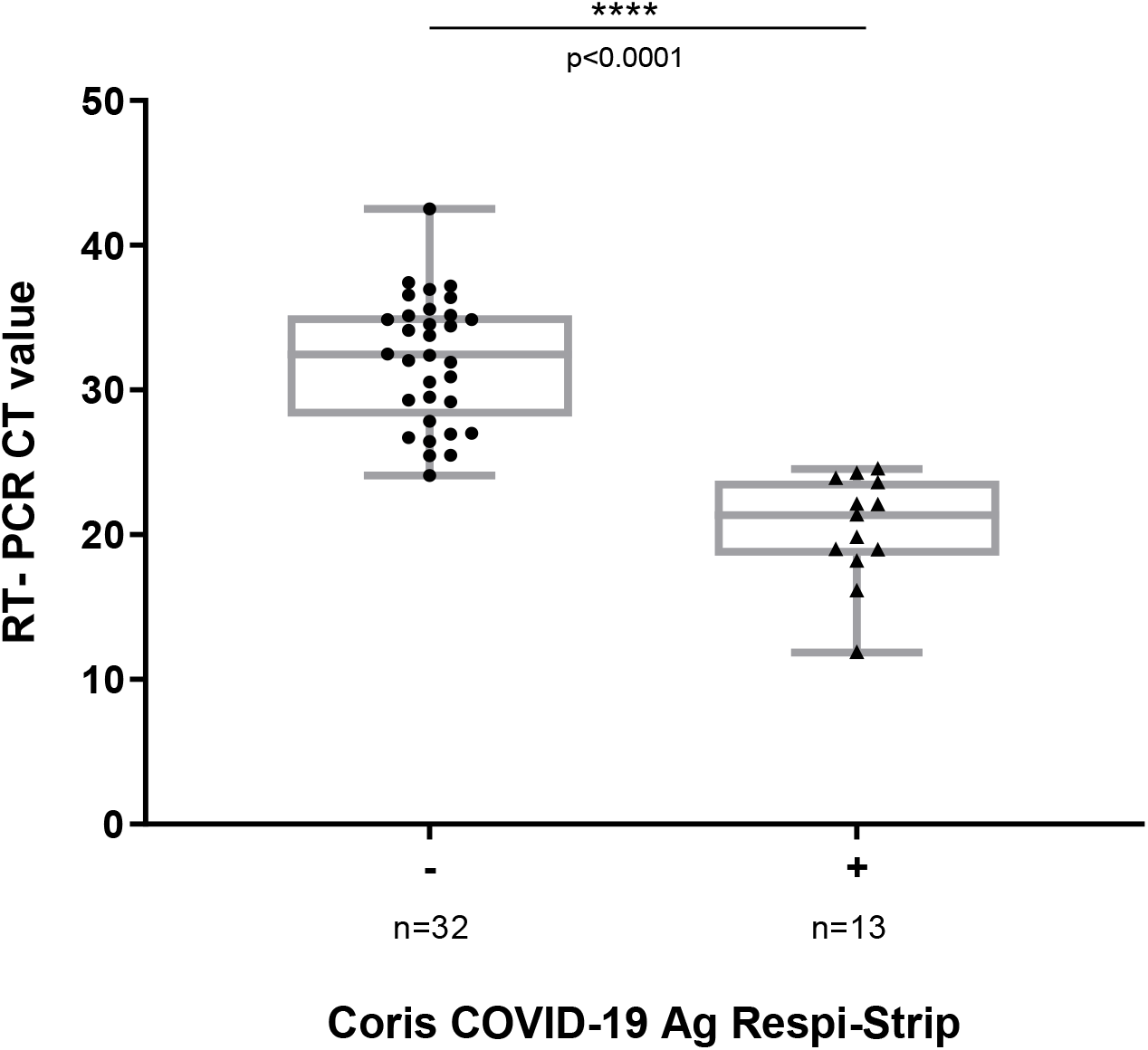
Coris COVID-19 Ag Respi-Strip results according to RT-PCR CT values. Cycle threshold (CT) values recorded by RT-PCR methods Allplex™ 2019-nCoV Assay Seegene. The boxes represent interquartile ranges with the horizontal line indicating the median CT value and the whiskers showing minimal and maximal CT values. The *p* value was calculated using the Mann-Whitney U test, and compares the distribution of CT values in serological diagnosis tests with positive and negative results.

Among the 45 blood samples collected in confirmed COVID-19 cases, 31 tested positive for IgM using the SureScreen RDT, resulting in a sensitivity (95% CI) of 68.9% (55.4 - 82.4).

Seventeen samples tested positive for IgM but negative for IgG using the serology RDTs. Among these samples, IgM were also detected by ELISA in 11 out of 17 cases (Sup. Fig S4. B). RT-PCR CT values were higher in patients who tested positive using the SureScreen IgM compared to those who tested negative (median (IQR): 31.9 (5.5 - 35.1) versus 25.0 (18.8 – 29.0), p=0.0084, Fig. 4). Hence, the sensitivity (95% CI) of the SureScreen IgM RDT was 46.7% (21.5 – 72.0) for CT values ≤ 25, reaching 86.7% (69.5 – 100.0) for CT values ≥ 35. Additionally, the SureScreen IgM RDT has a sensitivity (95% CI) of 64.0% (43.9 – 84.1) when the estimated time from the onset of symptoms was ≤ 7 days, reaching 78.0% (50.9 – 100.0) when ≥ 14 days (Sup. Table S1. A and B). Using the Szybio RDT, a total of 29 specimens tested positive for IgM, resulting in a sensitivity of 64.4% (50.5 – 78.4). Seventeen samples were IgM positive but IgG negative, and among these, IgM were also detected by ELISA in 11 samples (Sup. Fig S4. B). The CT value of the RT-PCR was higher in patients who tested positive for IgM using the Szybio RDT compared those who tested negative (median (IQR): 31.9 (24.9 – 35.0) versus 26.0 (19.6 – 31.3), p=0.0304, Fig. 4). The sensitivity of the Szybio IgM RDT was 46.7% (21.5 – 72.0) for CT values ≤ 25, and reached 80.0% (59.8 – 100.0) for CT values ≥ 35. In addition, the Szybio RDT had a sensitivity of 64.0 (43.9 – 84.1) when the estimated time from the onset of symptoms ≤ 7 days, reaching 67.0% (36.3 – 97.7) when ≥ 14 days (Sup. Tables S1. A and B).

All samples that tested positive for IgG using both the SureScreen and the Szybio RDTs also tested positive for IgM. The proportion of IgG positive samples rises according to the CT values and the time from the onset of COVID-19 symptoms (Sup. Tables S1. A and B).

### COVID-19 diagnosis using the antigen and IgG/IgM-based RDTs

The combination of the Coris COVID-19 Ag and the SureScreen IgM RDTs detected 38 out of 45 specimens, resulting in a sensitivity (95% CI) of 84.0% (73.3 – 94.7), while the combination of the Coris COVID-19 Ag and the Szybio IgM RDTs detected 36 specimens, resulting in a sensitivity of 80.0% (68.3 – 91.7) (Fig. 6). The combination of RDTs based on Ag plus IgM detection significantly improved the identification of COVID-19 cases at hospital admission compared to the Coris Ag RDT alone (p<0.0001) or the IgM/IgG RDTs alone (p=0.0113 and p=0.0142, respectively, Fig. 6).

**Figure 6:**
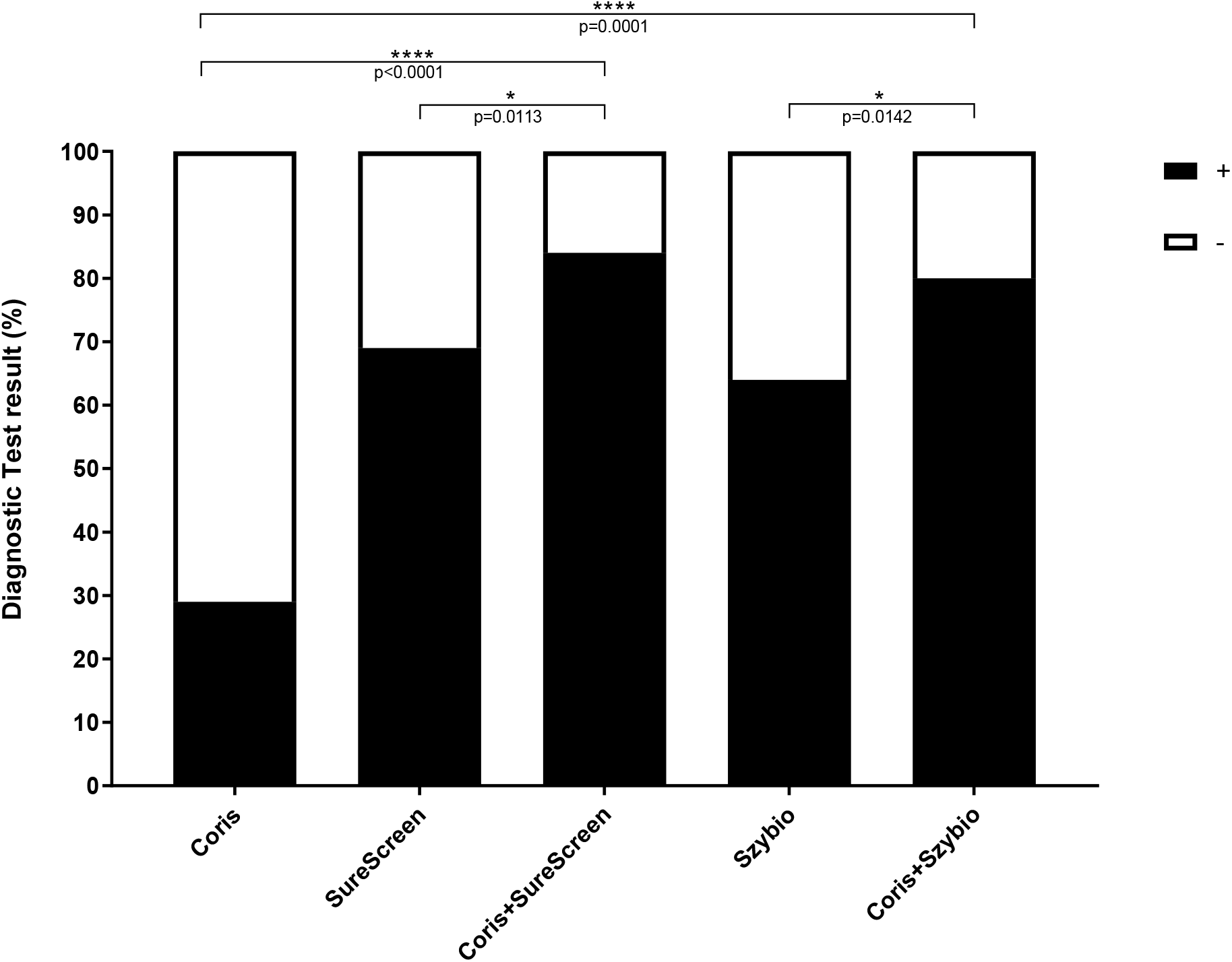
Performances of diagnostic strategies combining antigen and serological RDTs at hospital admission in patients with COVID-19 confirmed by RT-PCR. The diagnostic test results correspond to the proportion of patients with COVID-19 detected by RDTs. The *p* value was calculated using the Exact binomial’s test and compares the performances of a combination of antigen and serological RDTs versus antigen and serological RDTs use alone.

## IV. Discussion

In this study, we observed that a majority of nasopharyngeal samples with high concentrations of SARS-CoV-2 RNA collected in the early phase of the infection tested positive for COVID-19 using an antigen RDT, while IgM/IgG serological testing detected patients later in the course of the disease. A combination of an antigen-based RDT that indicates the presence of the virus, and an IgM-based RDT that determines whether or not a person has recently been immunized against the virus, may be useful in settings where access to fast RT-PCR methods is limited.

Our results using the Ag RDT on samples with high SARS-CoV-2 RNA concentrations are in line with previous studies that have reported sensitivities of 74.2% ^11^, 85.7% ^13^, and 100% ^12^ for CT values below 25. For specimens which tested positive with CT values over 25, the Ag RDT was not optimal because it was positive in only 12.5% of cases on a nasopharyngeal swab ^11^. Furthermore, according to Cochrane Library, the average sensitivity (95% CI) of the point-of-care antigen test corresponds to 56.2% (29.5 – 79.8) ^18^. A. Scohy *et al*. have estimated the lower limit of detection of the Coris Ag RDT to be equivalent to 1.8 × 10^5^ copies/mL ^12^. Success in SARS-CoV-2 isolation in nasopharyngeal specimens suggests that infectivity is low at this level of CT values ^4,19,20^. Data on the kinetics of SARS-CoV-2 RNA indicate that the viral load peaks within the first days after the onset of symptoms ^4^. Hence, Ag RDTs may be interesting in the early phase of the infection when the viral load is high and the risk of SARS-CoV-2 transmission is at its maximum. SARS-CoV-2 Ag testing requires nasopharyngeal collection and strict procedures with personal protective equipment to prevent SARS-CoV-2 acquisition. Despite this drawback, our results show that Ag testing using the Coris RDT can detect the virus before the development of a serological response, and Ag tests might have some advantages when used as a triage test in a pandemic context. First, Ag testing is simple and can be performed in about 15 minutes after the collection of samples. Secondly, the Coris Ag RDT that targets the SARS-CoV and SARS-CoV-2 highly conserved nucleoprotein antigen does not cross-react with seasonal coronaviruses ^11^. Hence, the specificity of the assay appears close to 100% in all published studies ^11–13^. Thanks to its high specificity and positive predictive value, a positive result using the Coris RDT would make it possible to avoid or delay the RT-PCR test ^11^. The two serological RDTs evaluated in this study also showed a high specificity but a variable sensitivity according to antibody isotype, time from onset of symptoms and RT-PCR CT values. The performances of the RDTs to detect IgG was lower than that of the Abbott SARS-CoV-2 IgG assay. While these RDTs had a lower capacity to detect low IgG concentrations compared to the chemiluminescence immunoassay, both the SureScreen RDT and the Szybio RDT had a good capacity to detect IgM (Sup. Figures S4. A and B). Results of previous studies suggest that the performances of immunoassays to detect IgM in the early phase of infection varies considerably according to the methods used. The detection of anti-SARS-CoV-2 IgM have been observed several days before the detection of IgG ^21,22^, but others studies reported IgM seroconversion at the same time or after IgG ^23–27^. It has been established that during the B cell response, IgM cells are present before IgG class switching. However, during the course of COVID-19 infection, the peak of IgM may be low and delayed compared to that of IgG. Data from a recent study comparing COVID-19 RDTs confirm that the analytical sensitivity for IgM is highly variable depending on the assay whereas the detection of IgG is more homogeneous ^28^, hence early IgM detection is strongly dependent on the test used. Beside detection of seroconversion in the early phase of infection, assessment of SARS-CoV-2 IgM may be of interest to distinguish between a recent versus a later immunization since most COVID-19 cases become seronegative for IgM within two months after symptom onset ^29^.

The overall efficiency of diagnostic strategies is not only characterized by the intrinsic performances of *in vitro* assays, which are mainly estimated through sensitivity and specificity, but also by their accessibility ^8^, effectiveness, speed of process, and period of time to obtain results. Furthermore, diagnosis can benefit from a testing algorithm based on successive steps of triage, screening, and confirmation. Tests performed at the point-of-care offer interesting benefits, including rapid diagnosis and simplicity of use outside of laboratory facilities ^18^. Our results have shown that a combination of antigen and antibodies-based RDTs is highly specific and detects most carriers of SARS-CoV-2 admitted to the hospital at different times over the course of the COVID-19 infection. A synthetic representation of variation over time in biological markers for COVID-19 diagnosis is proposed in Figure 7.

**Figure 7.**
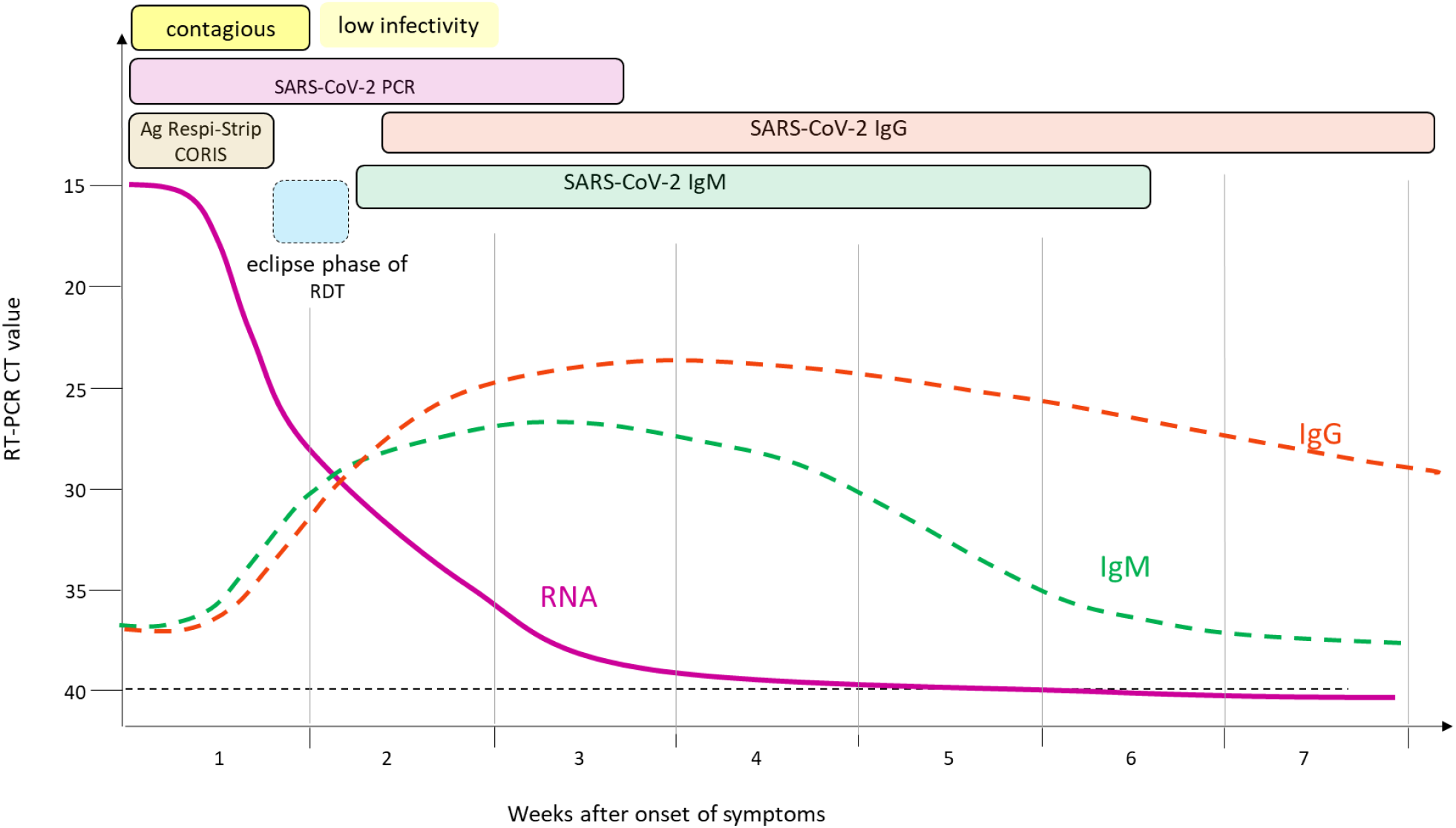
Variation over time in biological markers for COVID-19 diagnosis. High viral load (RT-PCR CT value < 25) and antigen detection in nasopharyngeal specimens characterize the first week after the onset of symptoms when the risk of SARS-CoV-2 transmission is at its maximum. The second week of COVID-19 infection is the period when the absence of detectable Ag and IgM/IgG is the most probable. The eclipse phase of antigen/IgM/IgG combined RDTs is most likely observable during this time period. A low level or the absence of SARS-CoV-2 RNA alongside IgG and IgM detection is observed two weeks after the onset of symptoms in most patients.

In conclusion, our results show that the Coris Ag immunochromatographic assay has insufficient sensitivity for the diagnosis of COVID-19 when used alone, but it could be a valuable tool when used in an integrative diagnostic strategy. Ag and IgM/IgG rapid diagnosis assays are complementary, and when used in combination are able to identify most patients with COVID-19 admitted in an emergency department. To be an effective alternative to nucleic acid tests and have a significant place in the global response to the COVID-19 pandemic, RDTs must combine high sensitivity to detect SARS-CoV-2 antigens and early and specific detection of IgM.

## Data Availability

Data will be obtainable after acceptance of the manuscript for publication through a simple request to the corresponding author.

## Notes

### Acknowledgments

We thank Grace Delobel for English language editing and review services.

## Supplemental data

**Supplemental Table S1. A:**
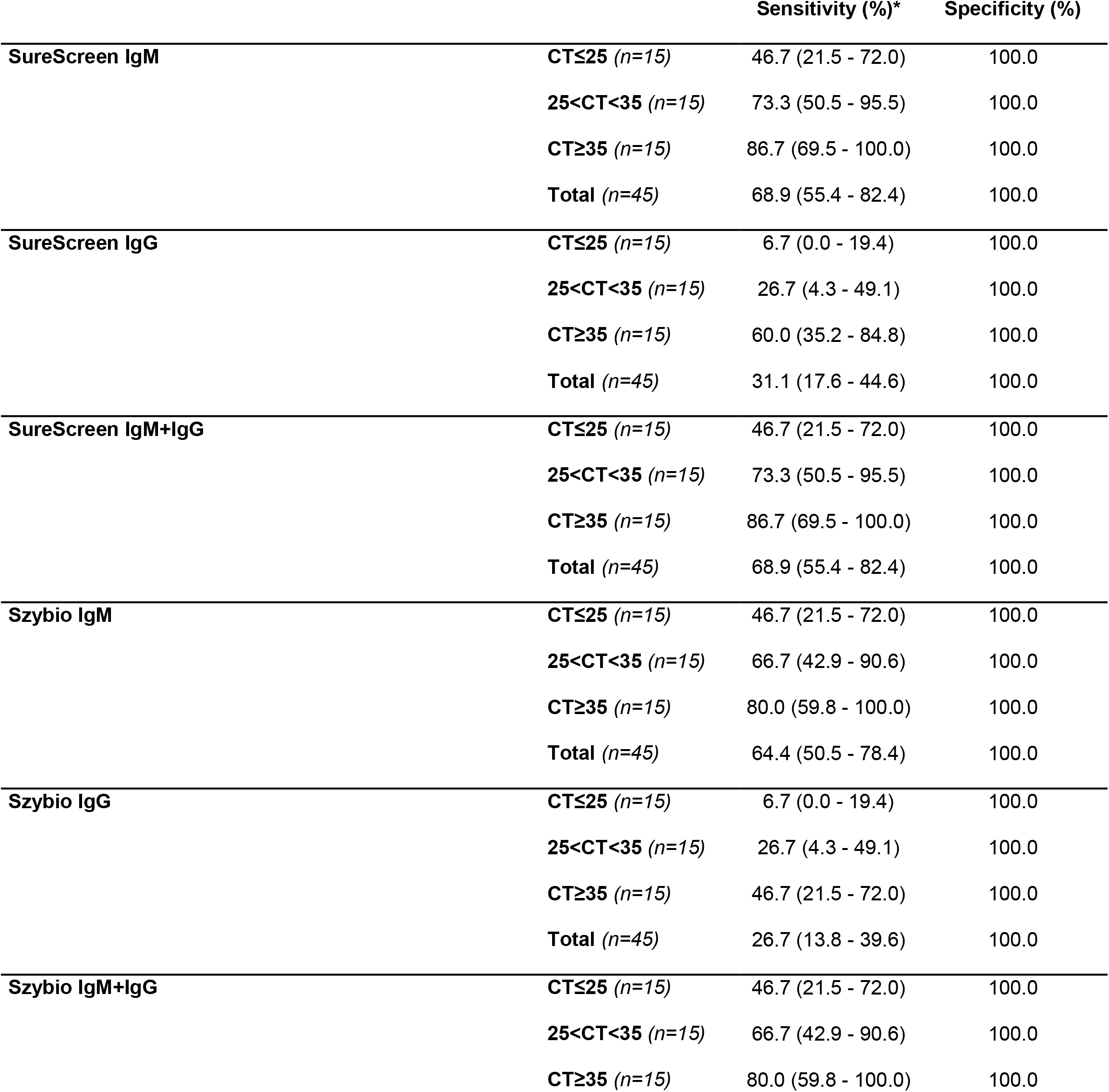
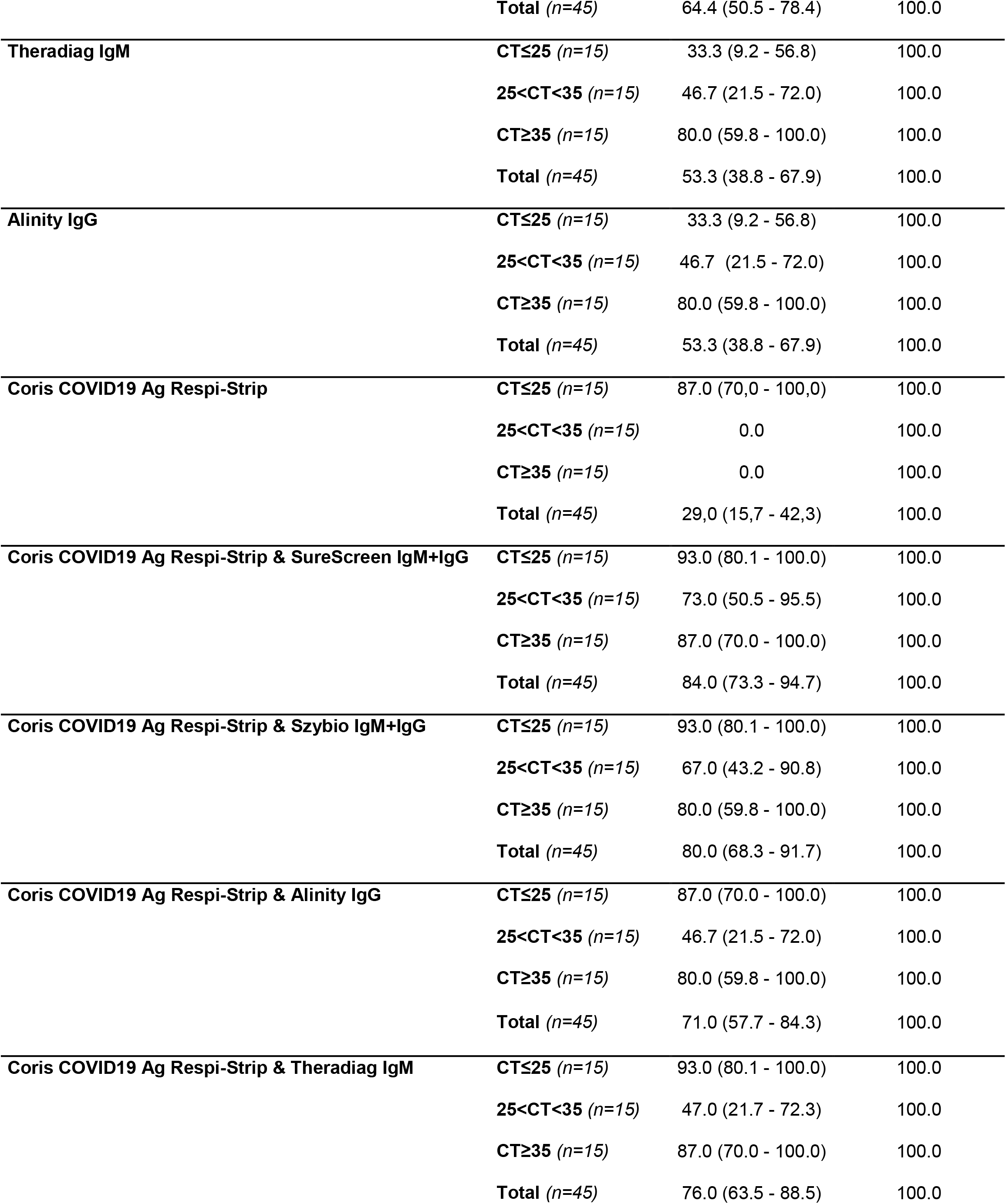
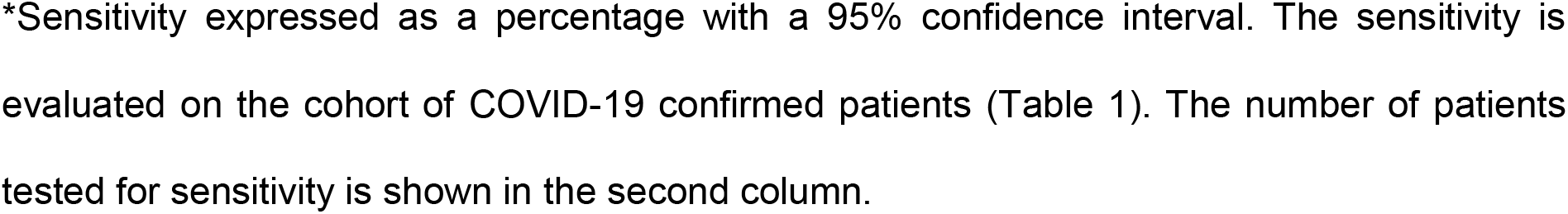
Performance of the assays observed in the study according to the RT-PCR CT value.

|  |  | **Sensitivity (%)\*** | **Specificity (%)** |
| --- | --- | --- | --- |
| **SureScreen IgM** | **CT≤25** (*n*=15) | 46.7 (21.5 - 72.0) | 100.0 |
|  | **25<CT<35** (*n*=15) | 73.3 (50.5 - 95.5) | 100.0 |
|  | **CT≥35** (*n*=15) | 86.7 (69.5 - 100.0) | 100.0 |
|  | **Total** (*n*=45) | 68.9 (55.4 - 82.4) | 100.0 |
| **SureScreen IgG** | **CT≤25** (*n*=15) | 6.7 (0.0 - 19.4) | 100.0 |
|  | **25<CT<35** (*n*=15) | 26.7 (4.3 - 49.1) | 100.0 |
|  | **CT≥35** (*n*=15) | 60.0 (35.2 - 84.8) | 100.0 |
|  | **Total** (*n*=45) | 31.1 (17.6 - 44.6) | 100.0 |
| **SureScreen IgM+IgG** | **CT≤25** (*n*=15) | 46.7 (21.5 - 72.0) | 100.0 |
|  | **25<CT<35** (*n*=15) | 73.3 (50.5 - 95.5) | 100.0 |
|  | **CT≥35** (*n*=15) | 86.7 (69.5 - 100.0) | 100.0 |
|  | **Total** (*n*=45) | 68.9 (55.4 - 82.4) | 100.0 |
| **Szybio IgM** | **CT≤25** (*n*=15) | 46.7 (21.5 - 72.0) | 100.0 |
|  | **25<CT<35** (*n*=15) | 66.7 (42.9 - 90.6) | 100.0 |
|  | **CT≥35** (*n*=15) | 80.0 (59.8 - 100.0) | 100.0 |
|  | **Total** (*n*=45) | 64.4 (50.5 - 78.4) | 100.0 |
| **Szybio IgG** | **CT≤25** (*n*=15) | 6.7 (0.0 - 19.4) | 100.0 |
|  | **25<CT<35** (*n*=15) | 26.7 (4.3 - 49.1) | 100.0 |
|  | **CT≥35** (*n*=15) | 46.7 (21.5 - 72.0) | 100.0 |
|  | **Total** (*n*=45) | 26.7 (13.8 - 39.6) | 100.0 |
| **Szybio IgM+IgG** | **CT≤25** (*n*=15) | 46.7 (21.5 - 72.0) | 100.0 |
|  | **25<CT<35** (*n*=15) | 66.7 (42.9 - 90.6) | 100.0 |
|  | **CT≥35** (*n*=15) | 80.0 (59.8 - 100.0) | 100.0 |
|  | **Total (n=45)** | 64.4 (50.5 - 78.4) | 100.0 |
| **Theradiag IgM** | **CT≤25 (n=15)** | 33.3 (9.2 - 56.8) | 100.0 |
|  | **25<CT<35 (n=15)** | 46.7 (21.5 - 72.0) | 100.0 |
|  | **CT≥35 (n=15)** | 80.0 (59.8 - 100.0) | 100.0 |
|  | **Total (n=45)** | 53.3 (38.8 - 67.9) | 100.0 |
| **Alinity IgG** | **CT≤25 (n=15)** | 33.3 (9.2 - 56.8) | 100.0 |
|  | **25<CT<35 (n=15)** | 46.7 (21.5 - 72.0) | 100.0 |
|  | **CT≥35 (n=15)** | 80.0 (59.8 - 100.0) | 100.0 |
|  | **Total (n=45)** | 53.3 (38.8 - 67.9) | 100.0 |
| **Coris COVID19 Ag Respi-Strip** | **CT≤25 (n=15)** | 87.0 (70.0 - 100.0) | 100.0 |
|  | **25<CT<35 (n=15)** | 0.0 | 100.0 |
|  | **CT≥35 (n=15)** | 0.0 | 100.0 |
|  | **Total (n=45)** | 29.0 (15.7 - 42.3) | 100.0 |
| **Coris COVID19 Ag Respi-Strip & SureScreen IgM+IgG** | **CT≤25 (n=15)** | 93.0 (80.1 - 100.0) | 100.0 |
|  | **25<CT<35 (n=15)** | 73.0 (50.5 - 95.5) | 100.0 |
|  | **CT≥35 (n=15)** | 87.0 (70.0 - 100.0) | 100.0 |
|  | **Total (n=45)** | 84.0 (73.3 - 94.7) | 100.0 |
| **Coris COVID19 Ag Respi-Strip & Szybio IgM+IgG** | **CT≤25 (n=15)** | 93.0 (80.1 - 100.0) | 100.0 |
|  | **25<CT<35 (n=15)** | 67.0 (43.2 - 90.8) | 100.0 |
|  | **CT≥35 (n=15)** | 80.0 (59.8 - 100.0) | 100.0 |
|  | **Total (n=45)** | 80.0 (68.3 - 91.7) | 100.0 |
| **Coris COVID19 Ag Respi-Strip & Alinity IgG** | **CT≤25 (n=15)** | 87.0 (70.0 - 100.0) | 100.0 |
|  | **25<CT<35 (n=15)** | 46.7 (21.5 - 72.0) | 100.0 |
|  | **CT≥35 (n=15)** | 80.0 (59.8 - 100.0) | 100.0 |
|  | **Total (n=45)** | 71.0 (57.7 - 84.3) | 100.0 |
| **Coris COVID19 Ag Respi-Strip & Theradiag IgM** | **CT≤25 (n=15)** | 93.0 (80.1 - 100.0) | 100.0 |
|  | **25<CT<35 (n=15)** | 47.0 (21.7 - 72.3) | 100.0 |
|  | **CT≥35 (n=15)** | 87.0 (70.0 - 100.0) | 100.0 |
|  | **Total (n=45)** | 76.0 (63.5 - 88.5) | 100.0 |

\*Sensitivity expressed as a percentage with a 95% confidence interval. The sensitivity is evaluated on the cohort of COVID-19 confirmed patients (Table 1). The number of patients tested for sensitivity is shown in the second column.

**Supplemental Table S1. B:**
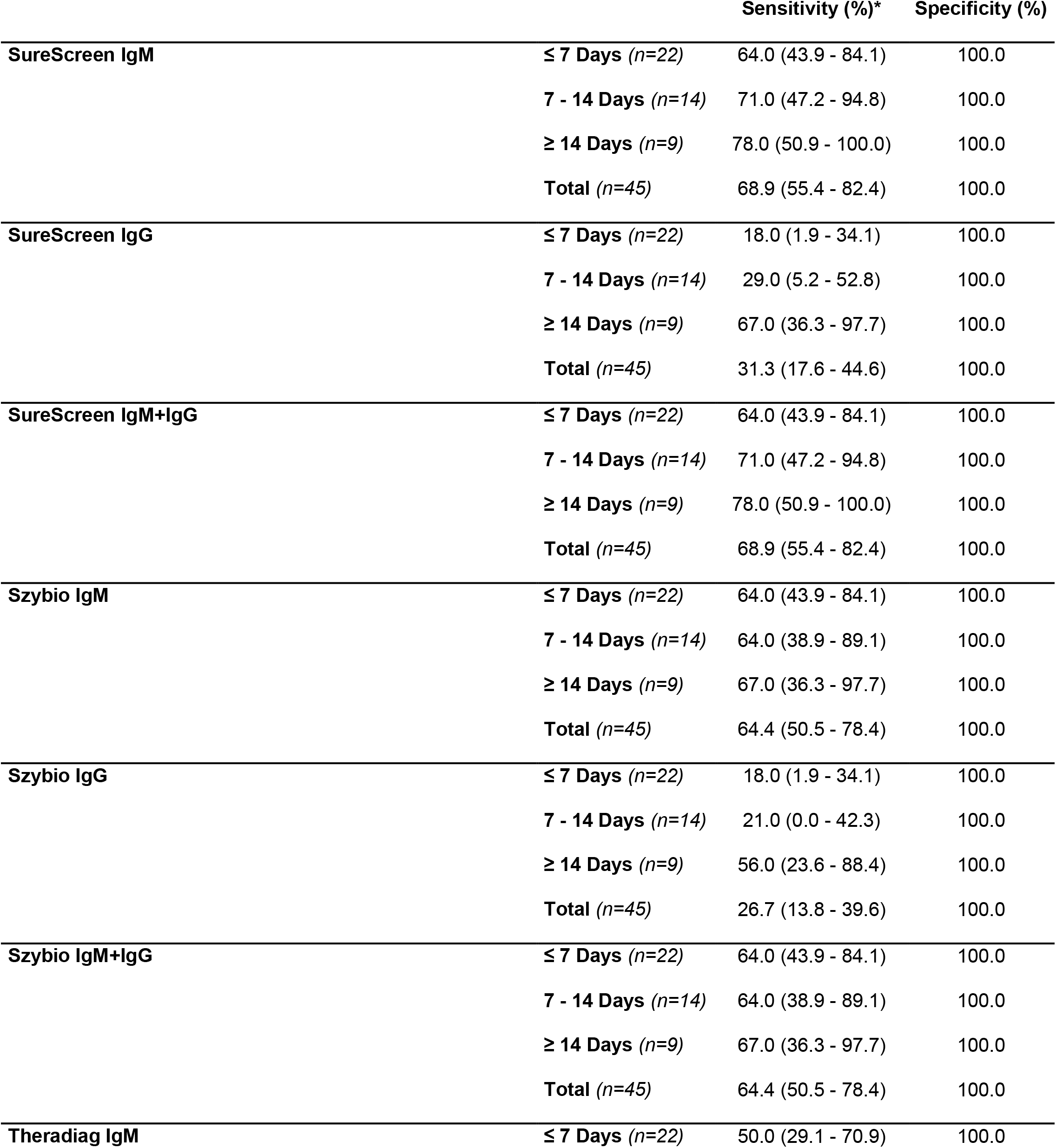
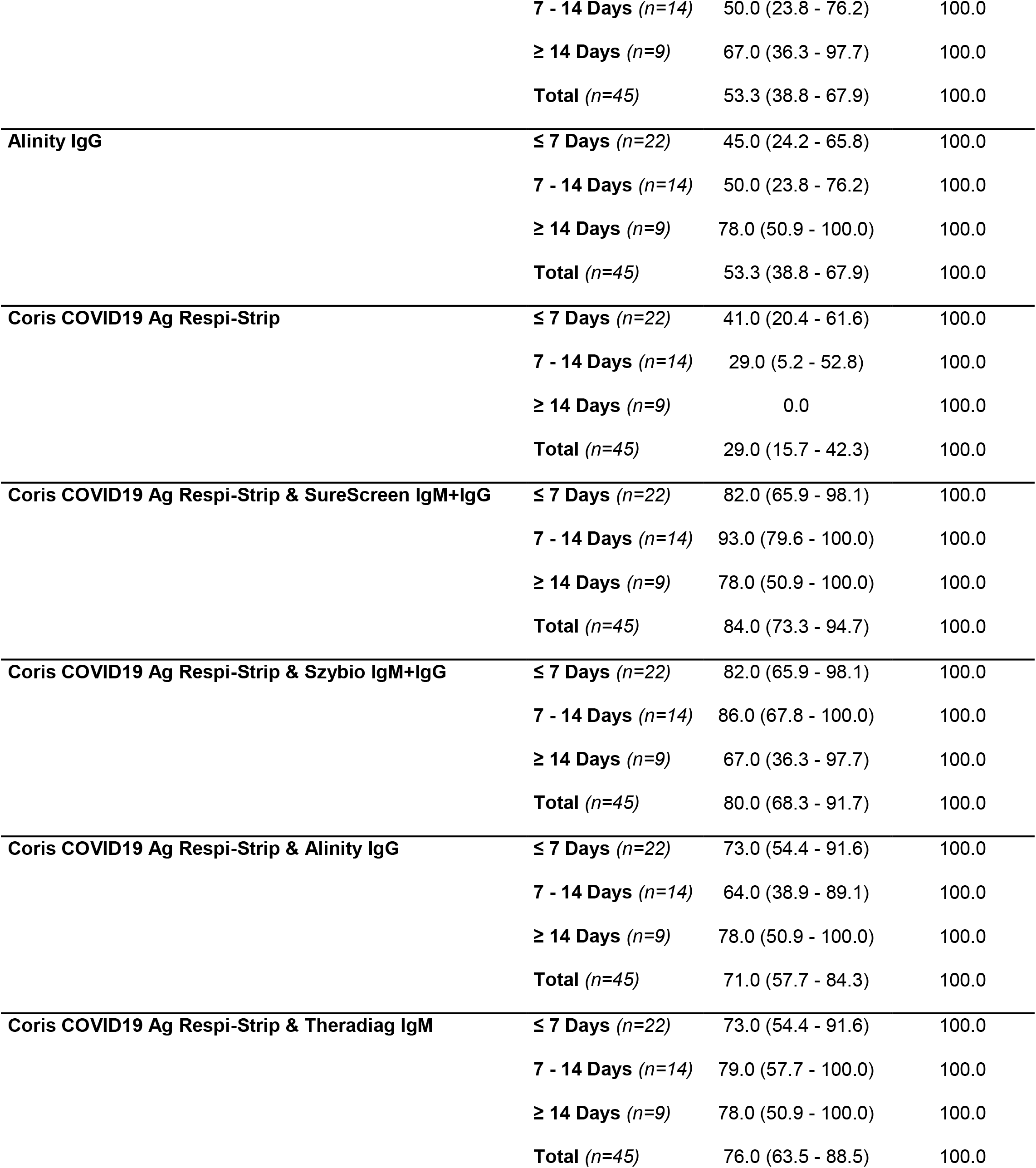
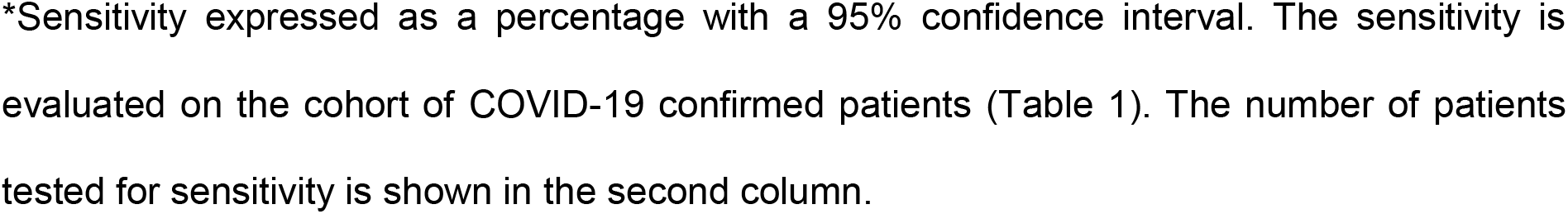
Performance of the assays observed in the study according to the time from onset of symptoms.

|  |  | **Sensitivity (%)\*** | **Specificity (%)** |
| --- | --- | --- | --- |
| **SureScreen IgM** | **≤ 7 Days (n=22)** | 64.0 (43.9 - 84.1) | 100.0 |
|  | **7 - 14 Days (n=14)** | 71.0 (47.2 - 94.8) | 100.0 |
|  | **≥ 14 Days (n=9)** | 78.0 (50.9 - 100.0) | 100.0 |
|  | **Total (n=45)** | 68.9 (55.4 - 82.4) | 100.0 |
| **SureScreen IgG** | **≤ 7 Days (n=22)** | 18.0 (1.9 - 34.1) | 100.0 |
|  | **7 - 14 Days (n=14)** | 29.0 (5.2 - 52.8) | 100.0 |
|  | **≥ 14 Days (n=9)** | 67.0 (36.3 - 97.7) | 100.0 |
|  | **Total (n=45)** | 31.3 (17.6 - 44.6) | 100.0 |
| **SureScreen IgM+IgG** | **≤ 7 Days (n=22)** | 64.0 (43.9 - 84.1) | 100.0 |
|  | **7 - 14 Days (n=14)** | 71.0 (47.2 - 94.8) | 100.0 |
|  | **≥ 14 Days (n=9)** | 78.0 (50.9 - 100.0) | 100.0 |
|  | **Total (n=45)** | 68.9 (55.4 - 82.4) | 100.0 |
| **Szybio IgM** | **≤ 7 Days (n=22)** | 64.0 (43.9 - 84.1) | 100.0 |
|  | **7 - 14 Days (n=14)** | 64.0 (38.9 - 89.1) | 100.0 |
|  | **≥ 14 Days (n=9)** | 67.0 (36.3 - 97.7) | 100.0 |
|  | **Total (n=45)** | 64.4 (50.5 - 78.4) | 100.0 |
| **Szybio IgG** | **≤ 7 Days (n=22)** | 18.0 (1.9 - 34.1) | 100.0 |
|  | **7 - 14 Days (n=14)** | 21.0 (0.0 - 42.3) | 100.0 |
|  | **≥ 14 Days (n=9)** | 56.0 (23.6 - 88.4) | 100.0 |
|  | **Total (n=45)** | 26.7 (13.8 - 39.6) | 100.0 |
| **Szybio IgM+IgG** | **≤ 7 Days (n=22)** | 64.0 (43.9 - 84.1) | 100.0 |
|  | **7 - 14 Days (n=14)** | 64.0 (38.9 - 89.1) | 100.0 |
|  | **≥ 14 Days (n=9)** | 67.0 (36.3 - 97.7) | 100.0 |
|  | **Total (n=45)** | 64.4 (50.5 - 78.4) | 100.0 |
| **Theradiag IgM** | **≤ 7 Days (n=22)** | 50.0 (29.1 - 70.9) | 100.0 |
|  | **7 - 14 Days** (n=14) | 50.0 (23.8 - 76.2) | 100.0 |
|  | **≥ 14 Days** (n=9) | 67.0 (36.3 - 97.7) | 100.0 |
|  | **Total** (n=45) | 53.3 (38.8 - 67.9) | 100.0 |
| **Alinity IgG** | **≤ 7 Days** (n=22) | 45.0 (24.2 - 65.8) | 100.0 |
|  | **7 - 14 Days** (n=14) | 50.0 (23.8 - 76.2) | 100.0 |
|  | **≥ 14 Days** (n=9) | 78.0 (50.9 - 100.0) | 100.0 |
|  | **Total** (n=45) | 53.3 (38.8 - 67.9) | 100.0 |
| **Coris COVID19 Ag Respi-Strip** | **≤ 7 Days** (n=22) | 41.0 (20.4 - 61.6) | 100.0 |
|  | **7 - 14 Days** (n=14) | 29.0 (5.2 - 52.8) | 100.0 |
|  | **≥ 14 Days** (n=9) | 0.0 | 100.0 |
|  | **Total** (n=45) | 29.0 (15.7 - 42.3) | 100.0 |
| **Coris COVID19 Ag Respi-Strip & SureScreen IgM+IgG** | **≤ 7 Days** (n=22) | 82.0 (65.9 - 98.1) | 100.0 |
|  | **7 - 14 Days** (n=14) | 93.0 (79.6 - 100.0) | 100.0 |
|  | **≥ 14 Days** (n=9) | 78.0 (50.9 - 100.0) | 100.0 |
|  | **Total** (n=45) | 84.0 (73.3 - 94.7) | 100.0 |
| **Coris COVID19 Ag Respi-Strip & Szybio IgM+IgG** | **≤ 7 Days** (n=22) | 82.0 (65.9 - 98.1) | 100.0 |
|  | **7 - 14 Days** (n=14) | 86.0 (67.8 - 100.0) | 100.0 |
|  | **≥ 14 Days** (n=9) | 67.0 (36.3 - 97.7) | 100.0 |
|  | **Total** (n=45) | 80.0 (68.3 - 91.7) | 100.0 |
| **Coris COVID19 Ag Respi-Strip & Alinity IgG** | **≤ 7 Days** (n=22) | 73.0 (54.4 - 91.6) | 100.0 |
|  | **7 - 14 Days** (n=14) | 64.0 (38.9 - 89.1) | 100.0 |
|  | **≥ 14 Days** (n=9) | 78.0 (50.9 - 100.0) | 100.0 |
|  | **Total** (n=45) | 71.0 (57.7 - 84.3) | 100.0 |
| **Coris COVID19 Ag Respi-Strip & Theradiag IgM** | **≤ 7 Days** (n=22) | 73.0 (54.4 - 91.6) | 100.0 |
|  | **7 - 14 Days** (n=14) | 79.0 (57.7 - 100.0) | 100.0 |
|  | **≥ 14 Days** (n=9) | 78.0 (50.9 - 100.0) | 100.0 |
|  | **Total** (n=45) | 76.0 (63.5 - 88.5) | 100.0 |

\*Sensitivity expressed as a percentage with a 95% confidence interval. The sensitivity is evaluated on the cohort of COVID-19 confirmed patients (Table 1). The number of patients tested for sensitivity is shown in the second column.

**Supplementa1 Figure S2:**
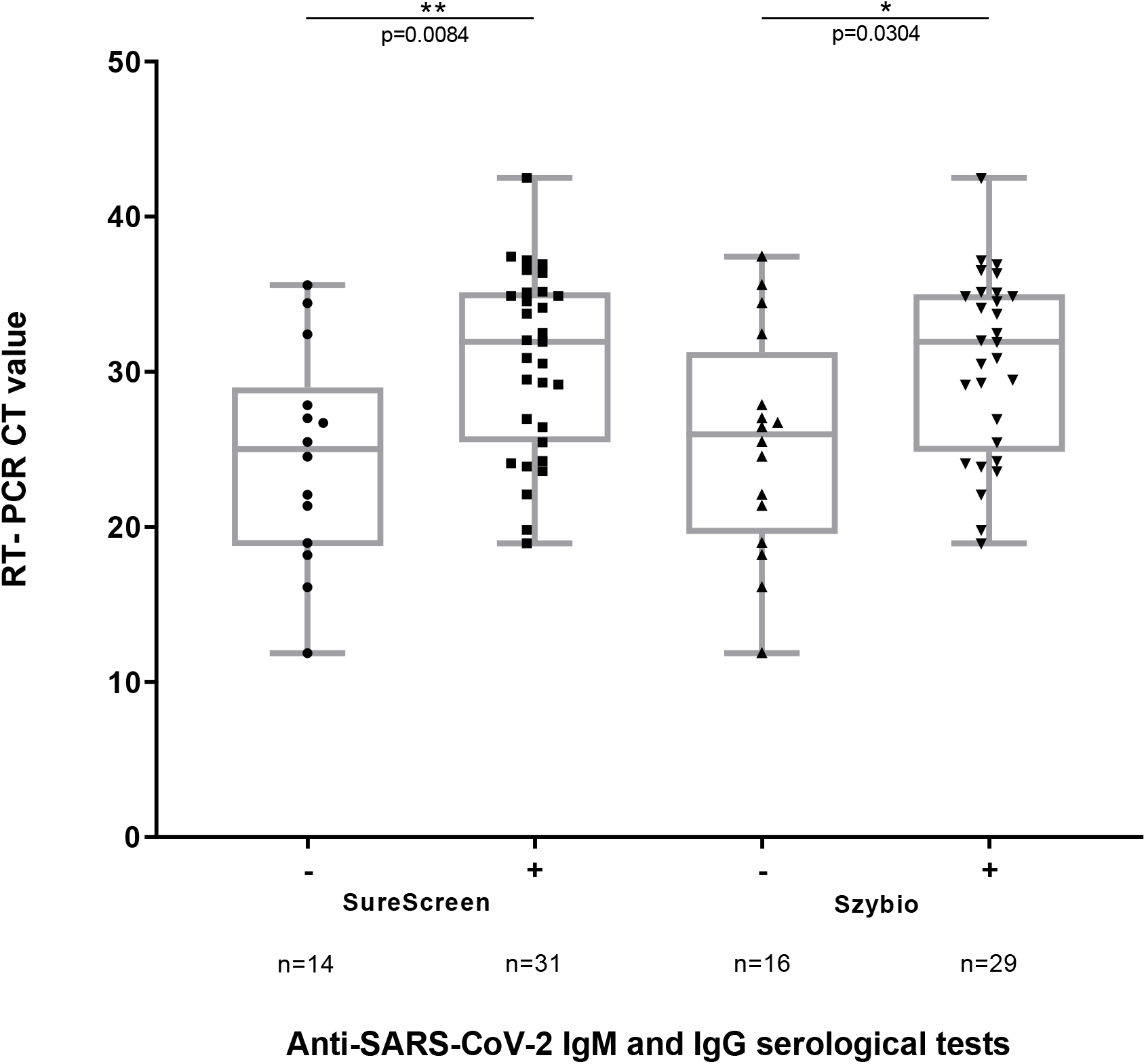
Anti-SARS-CoV-2 IgM and IgG detection using rapid serological diagnosis tests according to RT-PCR CT values. Cycle threshold (CT) values recorded by RT-PCR methods Allplex™ 2019-nCoV Assay Seegene. The boxes represent interquartile ranges with the horizontal line indicating the median CT values and the whiskers showing minimal and maximal CT values. The *p* value was calculated using the Mann-Whitney U test, and compares the distribution of CT values in serological diagnosis tests with positive and negative results.

**Supplemental Figure S3. A:**
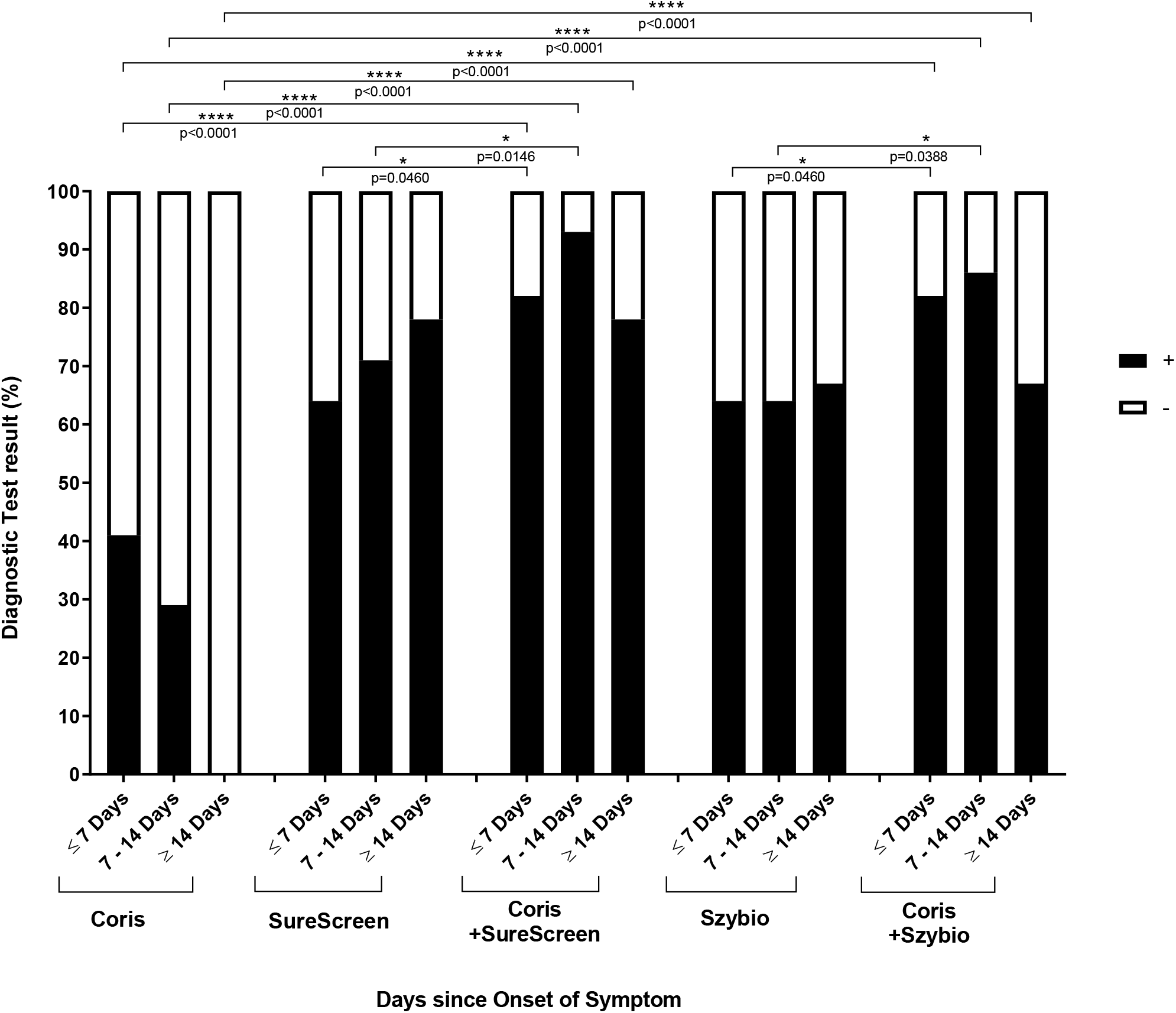
Diagnostic test results (%) according to test used and days since onset of symptoms. The diagnostic test results correspond to the proportion of patients with COVID-19 detected by RDTs. Patients in the cohort were divided into three groups according to the days since the onset of symptoms: ≤7 days (n=22); 7-14 days (n=14) and ≥14 days (n=9). The *p* value was calculated using Exact binomial’s test and compares, according to the days since the onset of symptoms, the performance of a combination of antigen and serological RDTs versus antigen and serological RDTs use alone.

**Supplemental Figure S3. B:**
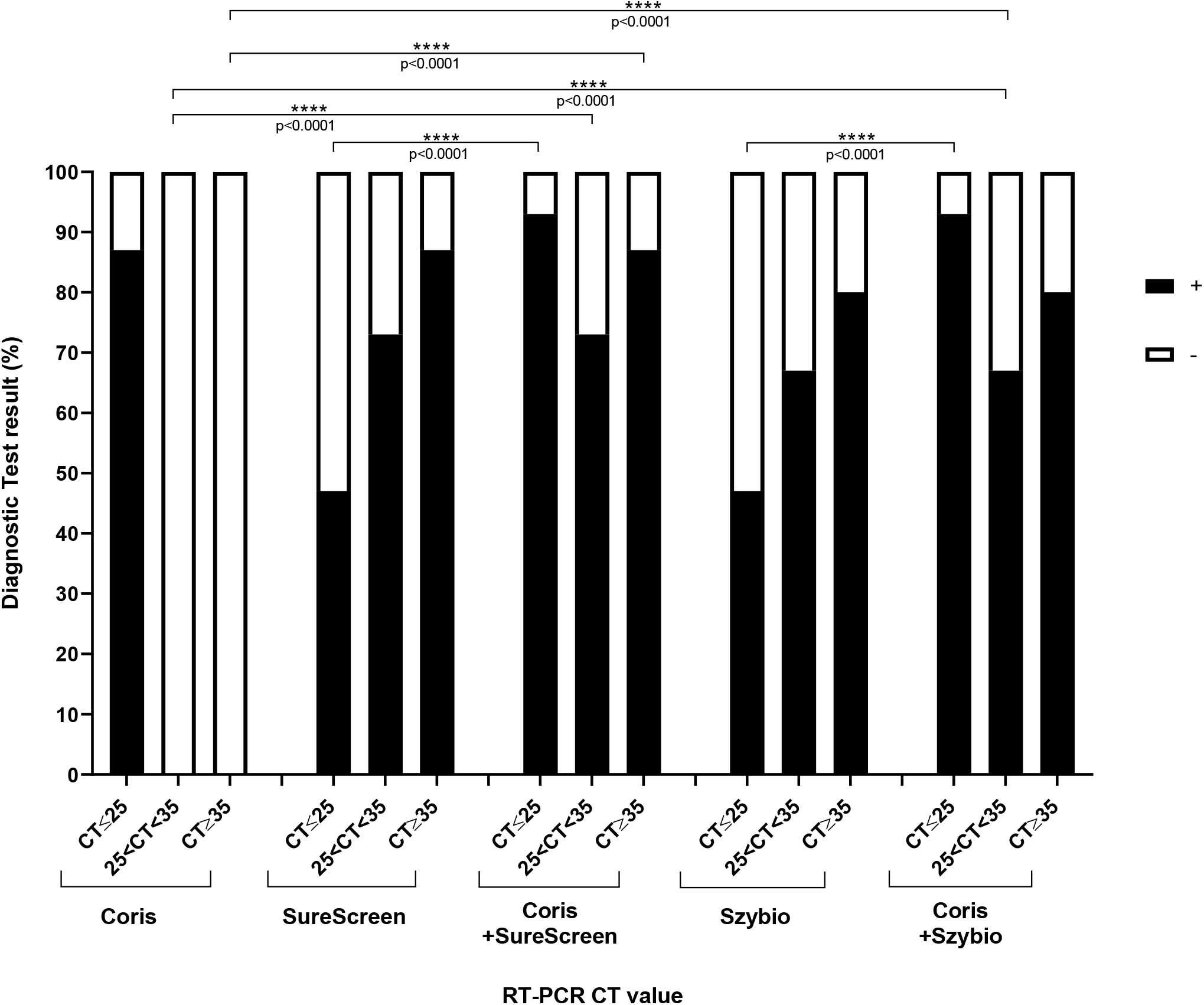
Diagnostic test results (%) according to test used and RT-PCR CT value. The diagnostic test results correspond to the proportion of patients with COVID-19 detected by RDTs. Patients in the cohort were divided into three groups according to RT-PCR CT values: CT≤25; 25<CT<35 and CT≥35. The *p* value was calculated using Exact binomial’s test and compares, according to the RT-PCR CT value, the performance of a combination of antigen and serological RDTs versus antigen and serological RDTs use alone.

**Supplemental Figure S4. A:**
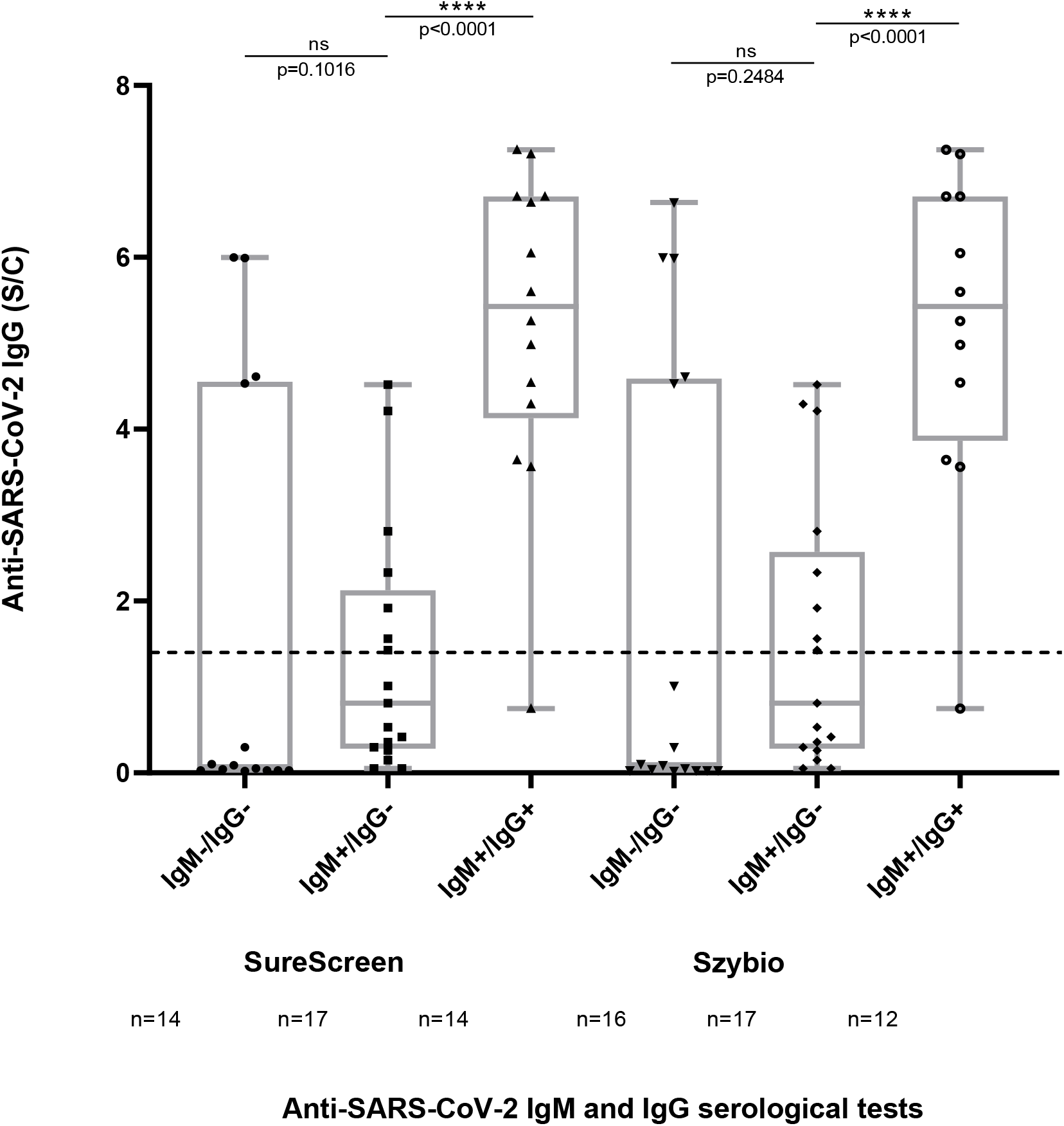
Anti-SARS-CoV-2 IgM & IgG detection using rapid serological diagnosis tests according to Abbott Alinity Anti-SARS-CoV-2 IgG (S/C). Anti-SARS-CoV-2 IgG serology was performed on the Alinity i Abbott automated system and index of the signal to control values (S/C) were reported. The dotted line indicates the positive threshold (≥ 1.4). The boxes represent interquartile ranges with the horizontal line indicating the median CT value and the whiskers showing minimal and maximal CT values. The *p* value was calculated using the Mann-Whitney U test and compares the distribution of IgG (S/C) Alinity i Abbott signals in serological diagnosis tests with IgM-/IgG-; IgM+/IgG- and IgM+/IgG+ results.

**Supplemental Figure S4. B:**
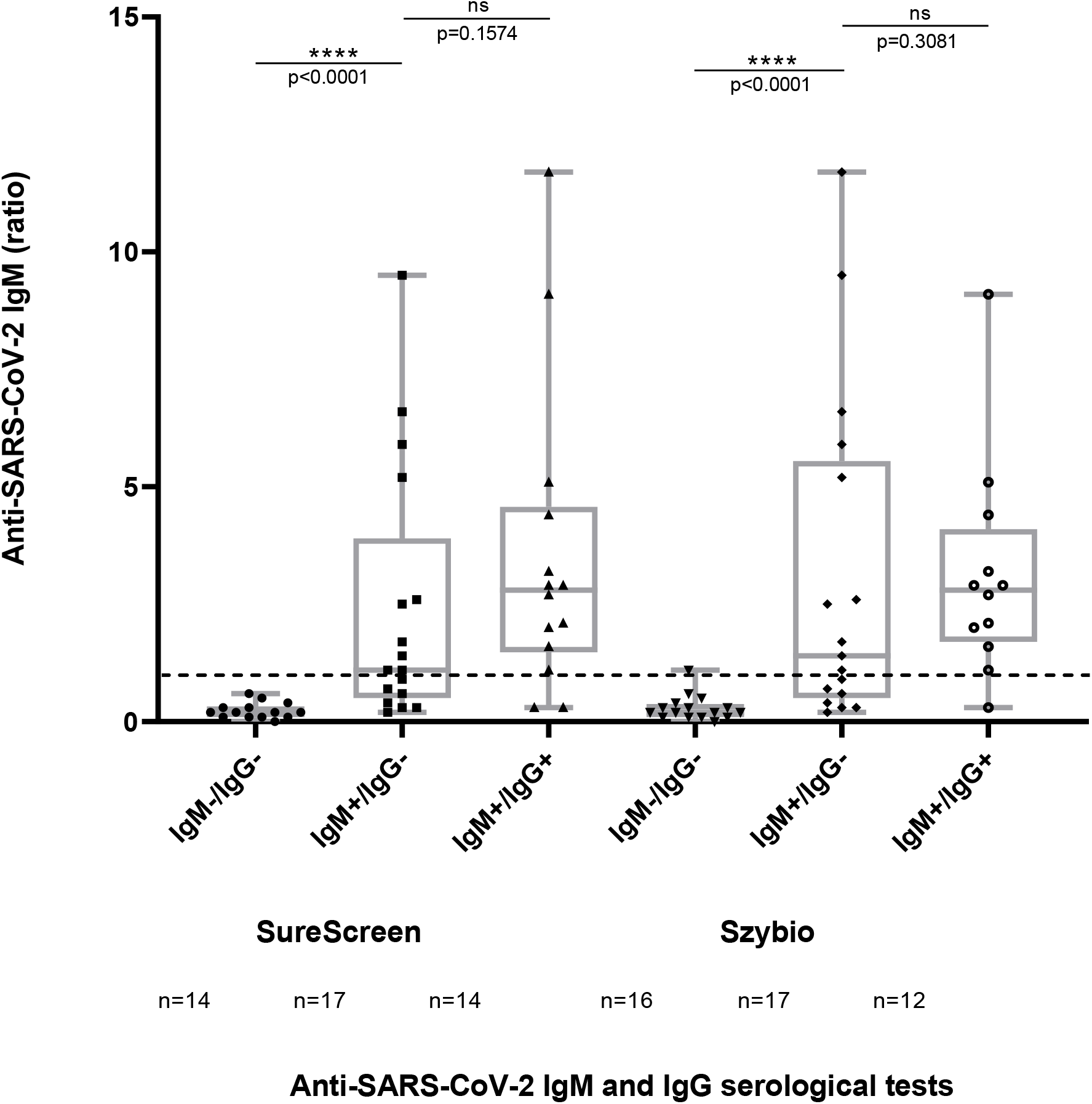
Anti-SARS-CoV-2 IgM & IgG detection using rapid serological diagnosis tests according to Theradiag ELISA Anti-SARS-CoV-2 IgM (ratio). Anti-SARS-CoV-2 IgM serology was performed by Theradiag ELISA test and the ratios of the signal were reported. The dotted line indicates the positive threshold (≥ 1.0). The boxes represent interquartile ranges with the horizontal line indicating the median CT value and the whiskers showing minimal and maximal CT values. The *p* value was calculated using the Mann-Whitney U test and compares the distribution of IgM Theradiag ELISA ratio in serological diagnosis tests with IgM-/IgG-; IgM+/IgG- and IgM+/IgG+ results.

